# Everolimus and mycophenolate mofetil effectively prevent GvHD in children with severe acute kidney injury undergoing allogeneic HSCT

**DOI:** 10.1101/2024.03.14.24304155

**Authors:** Felix Zirngibl, Pimrapat Gebert, Bianca Materne, Michael Launspach, Annette Künkele, Patrick Hundsdoerfer, Sandra Cyrull, Hedwig E Deubzer, Jörn-Sven Kühl, Angelika Eggert, Peter Lang, Lena Oevermann, Arend von Stackelberg, Johannes H Schulte

**Affiliations:** Charité – Universitätsmedizin Berlin, corporate member of Freie Universität Berlin, Humboldt – Universität zu Berlin, and Berlin Institute of Health, Department of Pediatric Oncology and Hematology, Berlin, Germany; Berlin Institute of Health at Charité – Universitätsmedizin Berlin, BIH Biomedical Innovation Academy, BIH Charité Clinician Scientist Program, Berlin, Germany; Charité – Universitätsmedizin Berlin, corporate member of Freie Universität Berlin, Humboldt – Universität zu Berlin, and Berlin Institute of Health, Institute of Biometry and Clinical Epidemiology, Berlin, Germany; German Cancer Consortium (DKTK), Heidelberg, Germany; German Cancer Research Center (DKFZ), Heidelberg, Germany; Department of Pediatrics, HELIOS Klinikum Berlin Buch, Berlin, Germany; Neuroblastoma Research Group, Experimental and Clinical Research Center (ECRC) of the Charité and the Max-Delbrück-Center for Molecular Medicine (MDC) in the Helmholtz Association, Berlin, Germany; Department of Pediatric Oncology, Hematology and Hemostaseology, University of Leipzig, Leipzig, Germany; Department of Hematology and Oncology, University Children’s Hospital, Eberhard Karls University Tuebingen, Tuebingen, Germany

## Abstract

Allogeneic hematopoietic stem cell transplantation (HSCT) serves as a therapeutic intervention for various pediatric diseases. Acute kidney injury afflicts 21-84% of pediatric HSCT cases, significantly compromising clinical outcomes. This retrospective single-institution analysis scrutinized the practice of substituting nephrotoxic ciclosporin A with the everolimus/mycophenolate mofetil combination as graft-versus-host disease (GVHD) prophylaxis in 57 patients following first allogeneic matched donor HSCT. The control cohort comprised 74 patients not receiving everolimus during the same timeframe. Study endpoints encompassed the emergence of retention parameters subsequent to the switch to everolimus, overall survival, relapse incidence of the underlying disease and acute and chronic GVHD in both treatment groups. Our findings reveal a significant improvement in renal function, evidenced by reduced creatinine and cystatin C levels 14 days after ceasing ciclosporin A and initiating everolimus treatment. Crucially, the transition to everolimus did not adversely affect overall survival post-HSCT (HR 1.4; 95% CI: 0.64 – 3.1; p=0.39). Comparable incidences of grade 2-4 and grade 3-4 acute GVHD as well as severe chronic GVHD were observed in both groups. Patients with an underlying malignant disease exhibited similar event-free survival in both treatment arms (HR 0.87, 95% CI: 0.39 – 1.9, p=0.73). This study provides compelling real-world clinical evidence supporting the feasibility of replacing CsA with everolimus and for the use of the everolimus/mycophenolate mofetil combination to manage acute kidney injury following HSCT in children.

**KEY POINTS:**

- Everolimus with or without MMF restores kidney function in children with acute kidney injury after allogeneic HSCT.
- Everolimus with or without MMF effectively prevent acute and chronic GvHD and leads to similar overall survival compared to standard therapy.

## INTRODUCTION

Allogeneic hematopoietic stem cell transplantation (HSCT) is a therapeutic option to treat malignant (leukemias and lymphomas) and non-malignant diseases (aplastic anemia, myelodysplastic syndrome, hemoglobinopathies, immunodeficiency disorders and inborn metabolic diseases).^1^ Recently, allogeneic HSCT has emerged as a a viable treatment approach for pediatric patients with solid tumors, such as neuroblastoma.^2^ Transplant related mortality (TRM) has demonstrated substantial improvement over the last decades. The 5-year overall survival (OS) rate rose from 41.8% in the period 1984-2001 to 79% in the years 2001 to 2009.^3^ Specifically, among children aged 4-18 years with a high-risk acute lymphoblastic leukemia in complete remission prior to first allogeneic HSCT, reported TRM rates fell below 10%.^4^ However, TRM is negatively influenced by patient age, donor type, and disease status^5,6^ and remains a substantial issue. Thus, it is crucial to further improve management of HSCT complications.

Acute kidney injury (AKI) occurs in 21-84% of pediatric HSCT cases in the literature and continues to be a severe problem.^7,8^ Patients requiring dialysis show a mortality rate of up to 77%.^9,10^ This association also applies to earlier-stage AKI as demonstrated by Kizilbash *et al*, who showed that a reduced OS correlates with an increasing severity of AKI.^8^ It remains therefore important, to prevent progression of AKI.

Biology of the underlying disease and infections, along with acute and chronic graft versus host disease (GVHD) are decisive determinants for the success of allogeneic HSCT.^11,12^ Patients experiencing advanced GVHD exhibit poorer OS.^13,14^ Consequently, GVHD prophylaxis is a primary challenge in clinical HSCT, particularly for patients with benign underlying diseases. A common strategy for GVHD prophylaxis involves a combination of ciclosporin A (CsA) and a short course of methotrexate (MTX), primarily used for myeloablative conditioning regimens. Nonmalignant diseases are often treated with a nonmyeloablative reduced intensity conditioning (RIC) regimen and a combination of CsA and mycophanolate mofetil (MMF).^15^ Additionally, serotherapy with anti-thymocyte globulin (ATG) is applied to further reduce the risk of acute and, especially, chronic GVHD.^16,17^ CsA is known to be a nephrotoxic drug, with nephrotoxicity attributed to vasoconstriction of the afferent arterioles.^18^ This vascular malfunction results from an increase in vasoconstrictive factors including thromboxane, endothelin, renin-angiotensin system activation, as well as a reduction of vasodilators like nitric oxide, prostacyclin and prostaglandin E2 (reviewed in *Naesens et al*^19^). Tacrolimus, another calcineurin inhibitor (CNI) besides CsA, less frequently used in pediatrics, also harbors nephrotoxic properties similar to those of CsA.^20,21^ Consequently, alternative immunosuppressive drug combinations must be found for patients with either preexisting or therapy-induced kidney injury.

Sirolimus (rapamycin), named after its discovery site, the Easter Island (*Rapa Nui*), is a naturally occurring compound isolated from a soil saprophyte. It inhibits the mammalian target of rapamycin (mTOR), an essential regulator of cell cycle in proliferating T cells. As an immunosuppressant, everolimus has been successfully used after solid organ transplantation in a combinatorial approach with CsA to prevent allograft rejection.^22^ Everolimus is a hydroxyethylester derivative of sirolimus with a shorter half-life (22 vs 72 hours), making everolimus serum levels better manageable in daily clinical practice. Orally bioavailable everolimus is rapidly absorbed, reaching maximum drug concentrations after 1 and 2 hours.^23^ Metabolization primarily occurs in the gut and liver by cytochrome P450 (CYP) 3A4, 3A5, and 2C8.^23^ Major class-effect toxicities in cancer patients were stomatitis, infections, noninfectious pneumonitis, fatigue, rash and diarrhea.^24^ On the other hand, everolimus lacks nephrotoxicity.^25^ Patients who converted early after kidney transplantation from CsA to everolimus showed greater improvement of renal function compared to CsA-treated controls.^26^ In this retrospective single center analysis we evaluated the practice at our institution to substitute CsA with the combination everolimus/MMF as GVHD-prophylaxis after first allogeneic HSCT in patients with severe AKI. To date, it is still unclear whether the immunosuppressive capacity of everolimus alone or in combination with MMF, without the use of calcineurin inhibitors, is sufficient to prevent GVHD, and so far, no studies exist to prove GVHD prophylaxis efficacy of everolimus in pediatric HSCT.

## PATIENTS AND METHODS

### Study design, setting and participants

This retrospective cohort study includes all patients treated with everolimus as GVHD prophylaxis after first allogeneic stem cell transplantation at Charité University Medicine Berlin between August 16, 2016 and September 29, 2020. The control cohort consisted of patients who underwent their first HSCT within the same timeframe but did not receive everolimus at any point post-transplantation. Patients who underwent mismatched family donor transplantation without subsequent CNI treatment for GVHD prophylaxis were excluded. This work only contains routinely acquired data, presented in an anonymous form. The study was ethically approved by the institutional review board, *Charité’s Ethics Committee,* under the reference EA2/144/15. This study adheres to the Strengthening the Reporting of Observational Studies in Epidemiology (STROBE) principles.

### Variables

The primary outcomes were incidence of acute and chronic GVHD, OS and relapse of the underlying malignant disease. Additional variables included in the analysis were demographic features, transplant type, conditioning type, plasma creatinine levels, plasma cystatin C levels and plasma everolimus levels.

### Data sources/ measurement

The medical records of all patients were evaluated for demographic features, dates of treatment and disease progression, received treatment, conditioning regimens used for HSCT, plasma creatinine and plasma cystatin C, occurrence of acute renal failure, everolimus plasma levels, occurrence of death, occurrence of acute and chronic GVHD and last follow-up. All data analyzing the incidence of acute or chronic GVHD were censored at the date of underlying disease relapse or death. Staging and grading of acute GvHD were based on Glucksberg *et al* in accordance with the European Society for Blood and Marrow Transplantation (EBMT) recommendations.^27^ Relapse was defined as the recurrence of the underlying malignant disease (morphologic, cytogenetic or molecular). The OS was defined where death from any cause was considered an event. The Jaffé-method was employed for creatinine measurement. Everolimus plasma levels were determined by enzyme-linked immunosorbent assay.

### Bias

The replacement of standard immunosuppression (CsA) with the combination everolimus/MMF was only performed in patients with AKI or with severe neurotoxicity. This fact represents a selection bias for patients with a significant complication in in their HSCT course.

### Statistical methods

Since a patient’s everolimus covariate status changes over time, the Simon-Makuch method^28^ was employed to depict the probability of OS and event-free survival (EFS).incidence of GVHD and relapse. The Simon-Makuch method generates survival curves for different levels of a time-dependent covariate. This method appropriately aligns the number of patients at risk as events (everolimus started yes/no) develop after HSCT. Cox proportional hazards analysis, incorporating a time-dependent covariate, was applied for OS and EFS. Cumulative incidence for competing events were conducted to evaluate the incidence of relapse and acute and chronic GVHD, using cause-specific approach. The effect of switch to everolimus as GVHD prophylaxis on survival was tested using the Mantel-Byar test. A 2-tailed P-value < 0.05 was considered statistically significant. Computations were performed using GraphPad prism (LaJolla, CA, USA), Stata IC15 (StataCorp, 2017, College Station, TX, USA) and the statistical software ‘EZR’ (Easy R), which is based on R and R commander.^29,30^

## RESULTS

### Patients’ characteristics

A total of 57 patients received everolimus with or without MMF during their clinical course after their first allogeneic HSCT as GVHD prophylaxis. Four patients (7.0%, #2, #20, #41 and #46) switched from Ciclosporin A to everolimus due to neurotoxicity. Patients #5 and #54 had an impaired renal function already before HSCT and did not receive CsA but started with everolimus/MMF. The remaining 51 (89.5%) patients received everolimus ± MMF because of escalating retention parameters and subsequent acute renal failure (**Figure 1**). The everolimus cohort is older compared to the control cohort, representing their higher susceptibility to develop AKI. The everolimus cohort comprises a diverse range of malignant (n=18, 31.6%) and nonmalignant (n=39, 68.4%) underlying diseases. The most frequent diagnoses were acute lymphoblastic leukemia (ALL) (n=15, 26.3%) and sickle cell disease (n=13, 22.8%). One ALL patient (#14) had trisomy 21. The median time of follow up was 36.9 months (range: 0.6-71.7). One patient (#42) is lost to follow up from day 201, but still alive. A cohort of 74 patients undergoing first allogeneic HSCT, receiving a standard calcineurin inhibitor-based GVHD prophylaxis and never received everolimus, served as a control. One patient with a malignant underlying disease of the CsA group was lost to follow up on day 211 without further information. Patients’ and control group’s characteristics are shown in **Table 1**.

**Figure 1.**
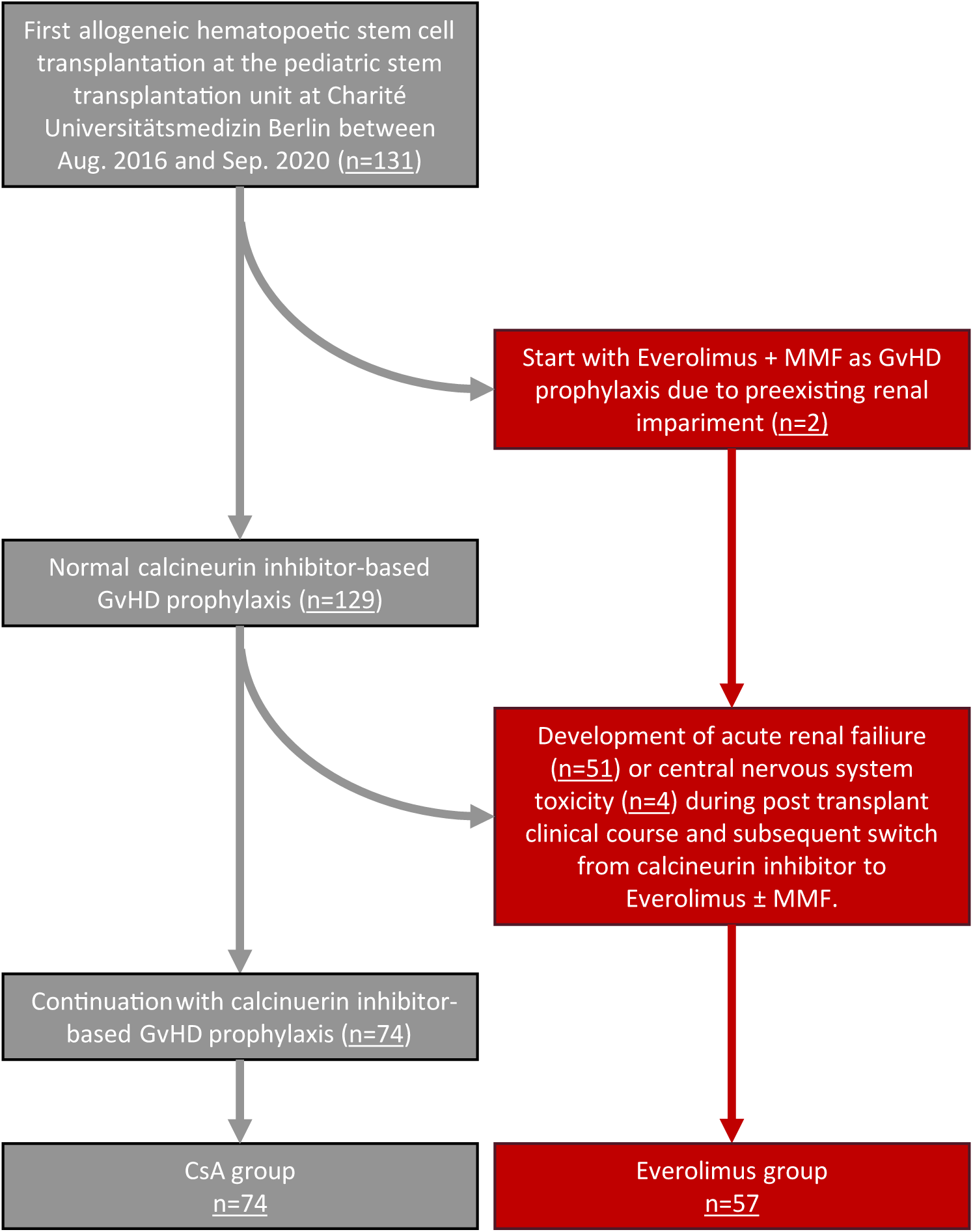
Study flow chart.

**TABLE 1.**
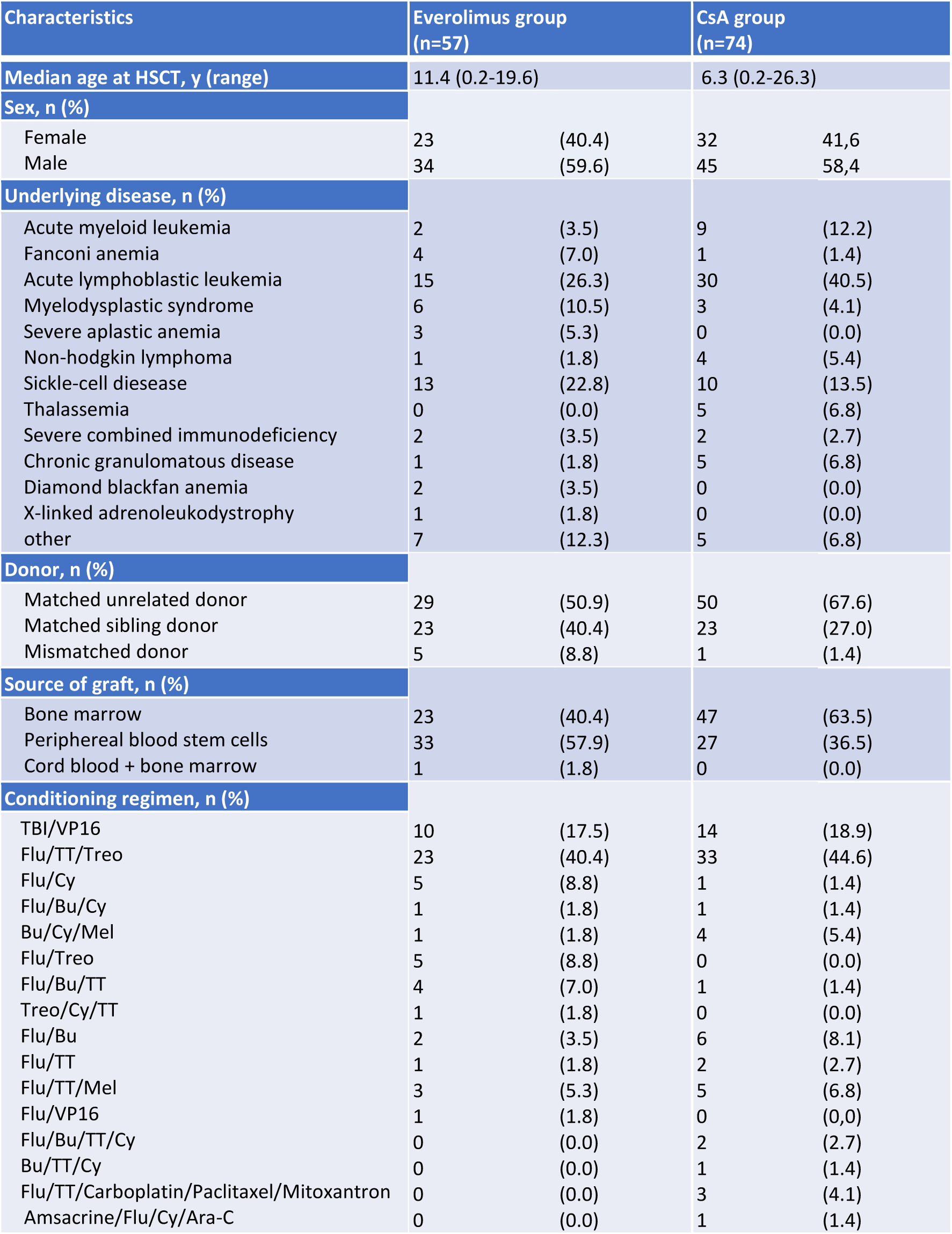
Patients’ characteristics.

### Descriptive Data

The median time of initiating Everolimus was 22 days (d) after HSCT ranging from –2 to 98. Median duration of everolimus treatment was 47 d (range: 11-128) for patients with a malignant underlying disease (#1-18) and 128 d (range: 11-355) for patients with a benign underlying disease (#19-57). The Median duration of MMF treatment was 29 d (range: 0-364) for patients with a malignant underlying disease (#1-18) and 64 d (range: 0-364) for patients with a benign underlying disease (#19-57). Everolimus was started at 1.6 mg/m²/d orally in two divided doses and dosing was subsequently adjusted to maintain blood concentrations between 3 and 8 ng/ml. Blood through levels for everolimus, measured by ELISA, are presented in **Figure 2** and were within the target range in most cases. If started, MMF was administered at a dose of 600 mg/m² twice daily. The immunosuppressive drugs used during the clinical course of the everolimus cohort is illustrated in **Figure 3**. Everolimus had to be discontinued in one patient who developed interstitial pneumonitis (#27) and in two patients who developed painful oral ulcers (#16, #56). Otherwise, we did not observe any toxicities necessitating discontinuation, including proteinuria, development of sinusoidal obstruction syndrome (SOS) or severe hepatotoxicity, although very high plasma levels were measured in some cases (**Figure 2**).

**Figure 2.**
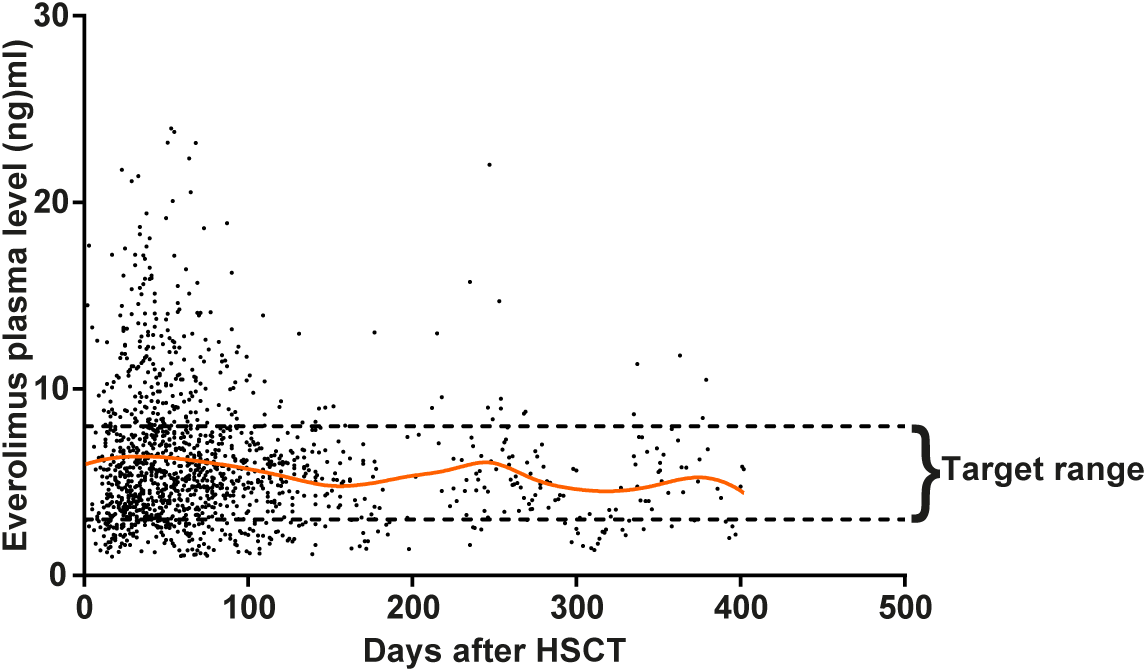
Everolimus plasma levels. The everolimus target ranges were 3 to 8 ng/ml (dashed lines). The solid line is a spline-smoothing curve.

**Figure 3.**
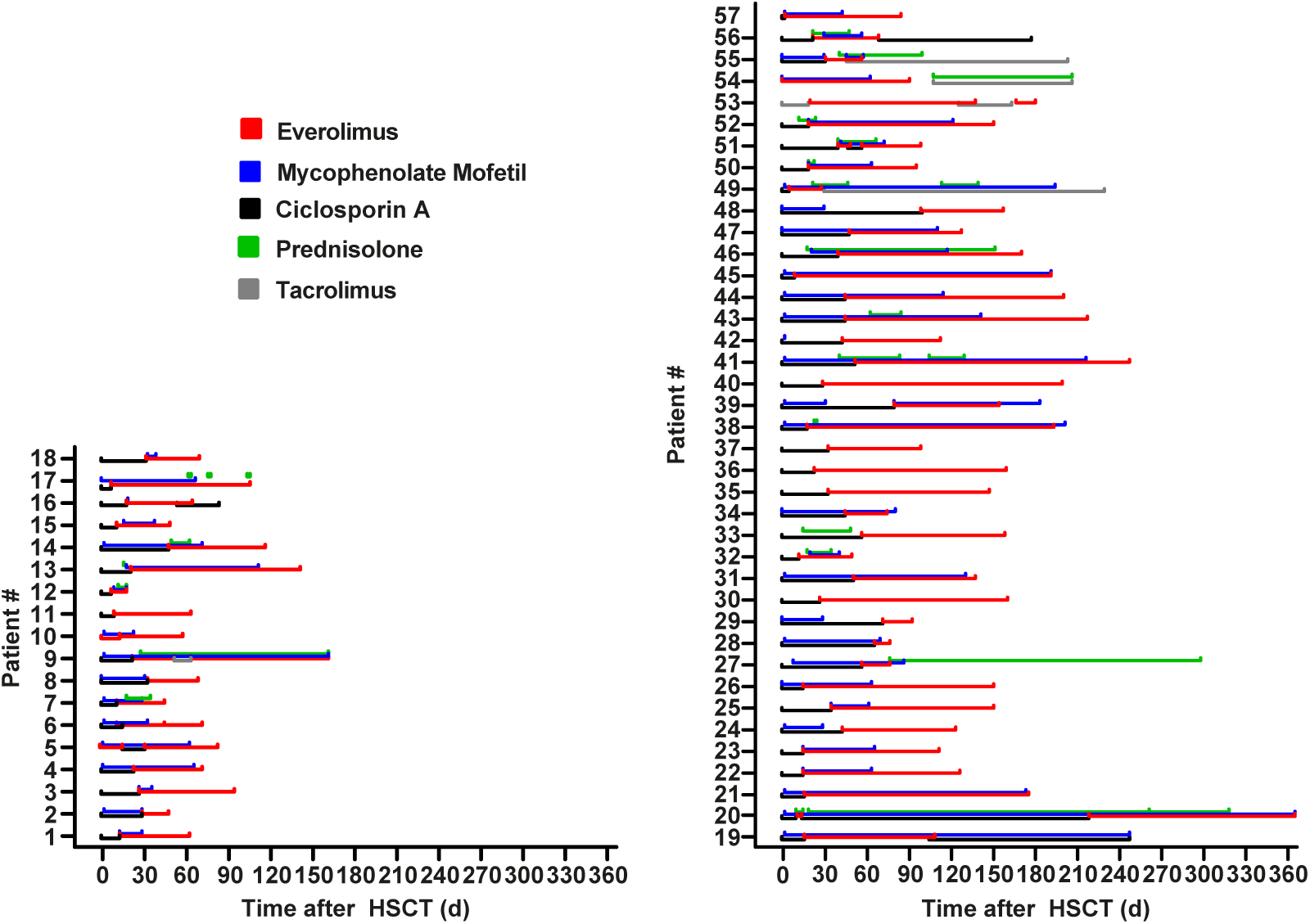
Timeline of immunosuppressive drugs used in patients that received everolimus ± mycophenolate mofetil. Each of the 57 pediatric patients that received everolimus at any time during their clinical course after hematopoetic stem cell transplantation is shown individually. Different colors indicate different immunosuppressive drugs. Patients #1 – 18 have malignant and patients #19 – 57 have benign underlying diseases.

### Renal function

Renal function was evaluated by measuring plasma creatinine (Jaffé-method) and plasma cystatin C relative to baseline at the day of transplantation. The decision to switch from standard CNI treatment to everolimus ± MMF was based on the development and the dynamics of acute renal failure and increasing retention parameters in the context of the patient’s general condition. When feasible, essential co-medication was dose-adjusted to the glomerular filtration rate or substituted with less nephrotoxic drugs, otherwise nephrotoxic medication was halted. For instance, amphotericin B was replaced by an azole and in some cases vancomycin was replaced by linezolid. If these interventions proved insufficient, ciclosporin A was stopped and everolimus was started. Everolimus plasma concentrations were monitored up to thrice a week, and the dose of everolimus was subsequently adjusted to achieve target trough levels (**Figure 2**). The median blood through levels were 5.46 ng/mL ranging from 1.01 to 23.97 ng/mL. After everolimus was started, plasma creatinine significantly decreased from a mean of 294 % (±158) relative to baseline, measured on the day of HSCT to 158 % (±67) 14d later (**Figure 4A**), plasma cystatin C decreased from 210 % (±68) of baseline to 135 % (±50) (**Figure 4B**). Patients #5 and #54 were excluded from this analysis as they initially started with everolimus. For patients #2, #14, #34, #40. #41, #42 and #46 no reasonable data for cystatin C were collected, leading to their exclusion from the analysis. Therefore, 55 patients could be included for creatinine analysis and 48 patients could be included for cystatin C analysis. Our data demonstrate that changing immunosuppression from CsA to everolimus with or without MMF, significantly improves renal function of pediatric patients post-HSCT.

**Figure 4.**
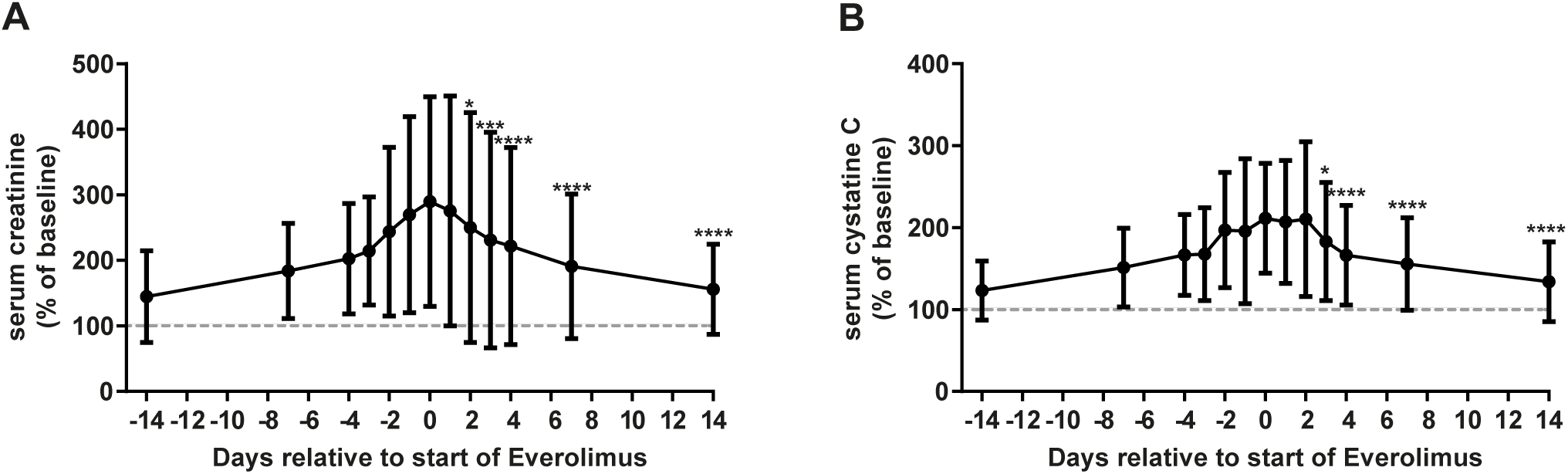
Development of retention parameters after the switch to everolimus ± mycophenolate mofetil (MMF). The grouped blood levels of creatinine **(A)** and cystatine C **(B)** of pediatric patients that developed acute renal failure during their post-HSCT course is shown at the time they were switched to the combination of everolimus ± MMF. Data is normalized to the creatinine and cystatine C level on the day of the HSCT, respectively, which is considered as baseline level. *: p<0.05, ***: p<0.001, ****: p<0.0001 using the mixed-effects model with Dunnet’s multiple comparisons test. HSCT: heamtopoetic stem cell transplantation.

### Graft versus host disease

Changing the immunosuppressive strategy during the post-HSCT course may result in the undesirable complication of acute or chronic GVHD. The cumulative incidence of competing events of grade II-IV aGVHD is shown in **Figure 5A**. In the everolimus group 10.0 % of patients develop grade II-IV aGVHD vs 18.2 % in the control cohort with a hazard ratio (HR) of 0.88 (95% CI: 0.29 – 2.7; p=0.82). The cumulative incidence of competing events of grade III-IV aGVHD is presented in **Figure 5A**. In the everolimus group 7.5 % of patients develop grade III-IV aGVHD, as opposed to 7.0 % in the control cohort with a HR of 1.82 (95% CI: 0.45 – 7.4; p=0.40) showing no significant difference. Consequently, patients that develop no signs of aGVHD until they switch to everolimus have a low risk of developing severe GVHD after this transition. Out of the everolimus cohort, 6 patients developed any signs of grade III-IV aGVHD, 3 (50 %) out of these patients had first GVHD symptoms before everolimus was started. Only one of the everolimus patients showed a relapse of mild GVHD symptoms (#14) after everolimus was started. Overall, most aGVHD symptoms in patients receiving everolimus were mild, and no severe relapses of preexisting GVHD were observed after the switch to everolimus. The incidence of severe chronic GVHD (cGVHD) was slightly higher in the everolimus cohort as depicted in **Figure 5C**. In the everolimus group 12.2 % of patients developed severe cGVHD and all had severe intestinal cGHVD, whereas 2 % of patients of the control cohort developed severe cGVHD (**Figure 5C**). This difference, however, was not statistically significant (HR 2.76, 95% CI: 0.69 – 11.0; p=0.15). Furthermore, we assessed possible confounders on GVHD incidence. Underlying diseases differed slightly in our study groups. However, ALL and sickle cell disease, the most prevalent underlying disease in our study, did not alter the HR of developing acute or chronic GVHD (**Table 2**). No events for grade 3-4 aGVHD were recorded for sickle cell disease in the everolimus group (**Table 2**). Furthermore, the graft source differed between the everolimus group and the CsA group. Patients who received everolimus were more frequently transplanted with a manipulated peripheral blood stem cell graft containing a defined amount of T-cells. Manipulated peripheral blood stem cell grafts did not lead to higher rates of acute or chronic GHVD compared to bone marrow grafts. Thus, we could rule out the graft source as a confounding variable.

**Figure 5.**
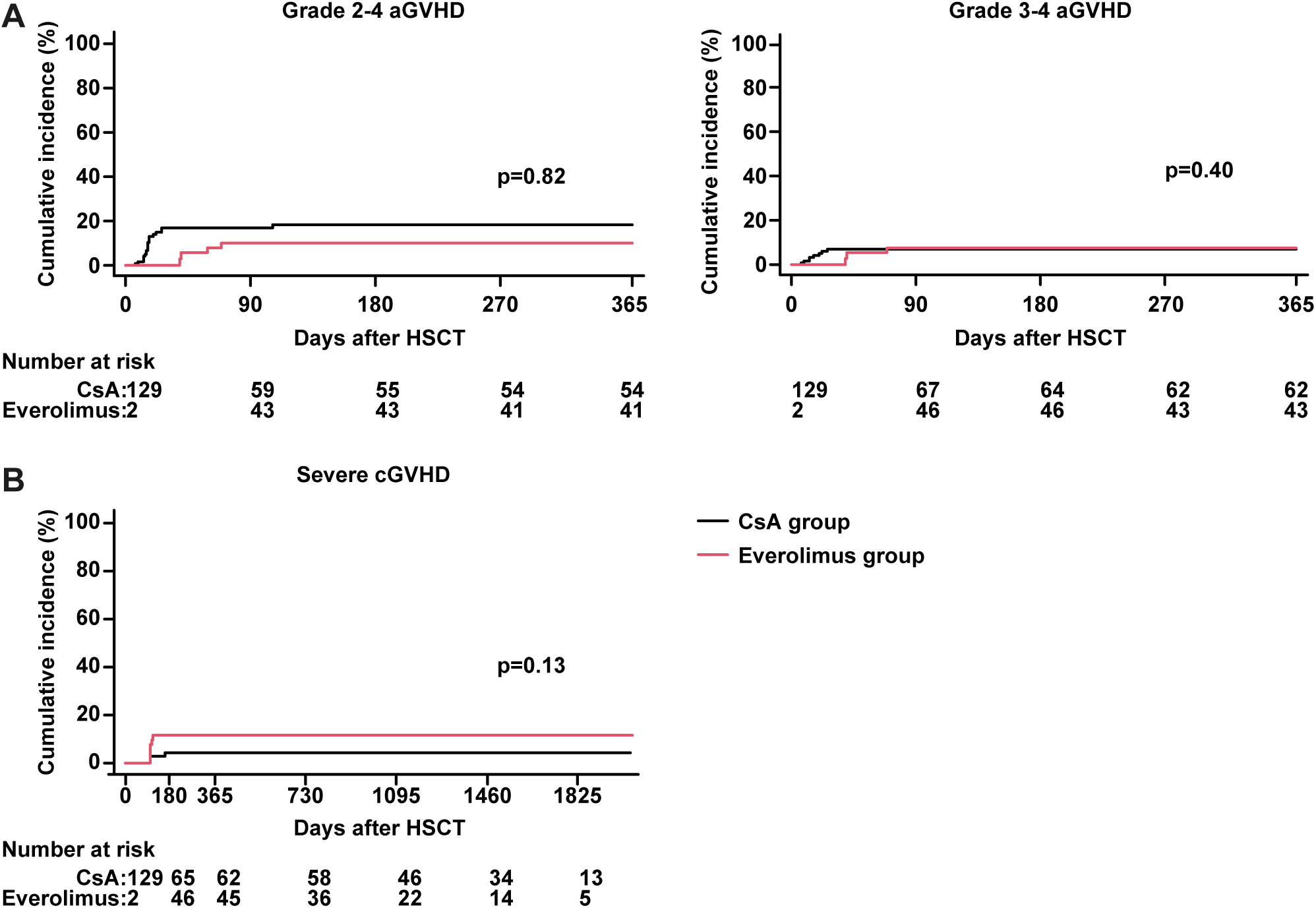
Cumulative incidence of acute and chronic graft versus host disease (GVHD). The cumulative incidence of **(A)** grade 2-4 acute (a)GVHD and grade 3-4 aGVHD and **(C)** severe chronic (c)GVHD is shown for all patients. Incidence curves are generated using a competing events model to reflect the time dependent covariate of switch to everolimus. P-value is determined using the Mantel-Byar test. CsA: Ciclosporin A

**TABLE 2.**
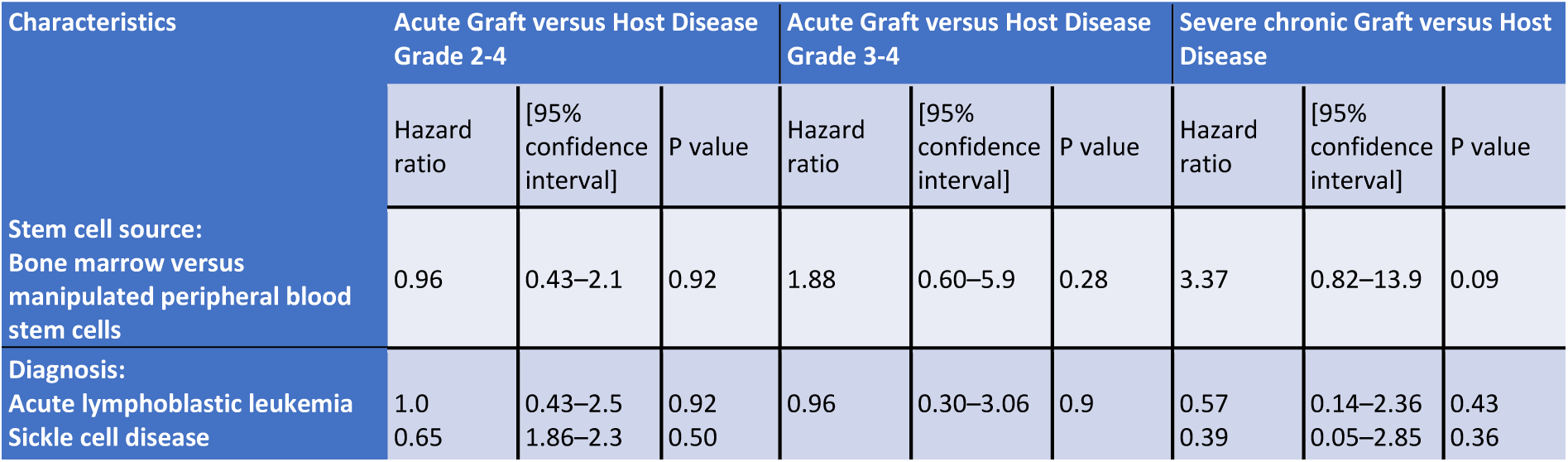
Adjusted Hazard ratios for graft source and underlying disease status “acute lymphoblastic leukemia”.

### Overall and event-free survival

The OS did not show a significant difference between the two groups (HR 1.4; 95% CI: 0.64 – 3.1; p=0.39). The 100-day and 2-year OS were 92.3% and 79.8% for the everolimus group and 95.3% and 84.1% in the control cohort, respectively (**Figure 6**). However, patients with a malignant underlying disease exhibited a significantly lower OS compared to the control cohort (HR 2.7, 95% CI: 1.1 – 6.9, P=0.04) (**Figure 7A**). The 2-year OS for this cohort was 58.6% in the everolimus group versus 83.7% in the control group. On the contrary, the relapse incidence after 2 years was higher in patients of the control cohort at 52.8% compared to 28.2% for everolimus patients (HR=0.55; 95% CI: 0.19 – 1.6; p=0.27), although this difference was not statistically significant (**Figure 7B**). Taken together, these findings result in an equivalent EFS (HR 0.84, 95% CI: 0.43 – 2.0, P=0.88) (**Figure 7C**). In conclusion, we demonstrate a non-inferiority in OS for patients receiving everolimus as GVHD prophylaxis, conversely, we observe a trend suggesting that everolimus might be beneficial for relapse-free survival in patients with a malignant disease.

**Figure 6.**
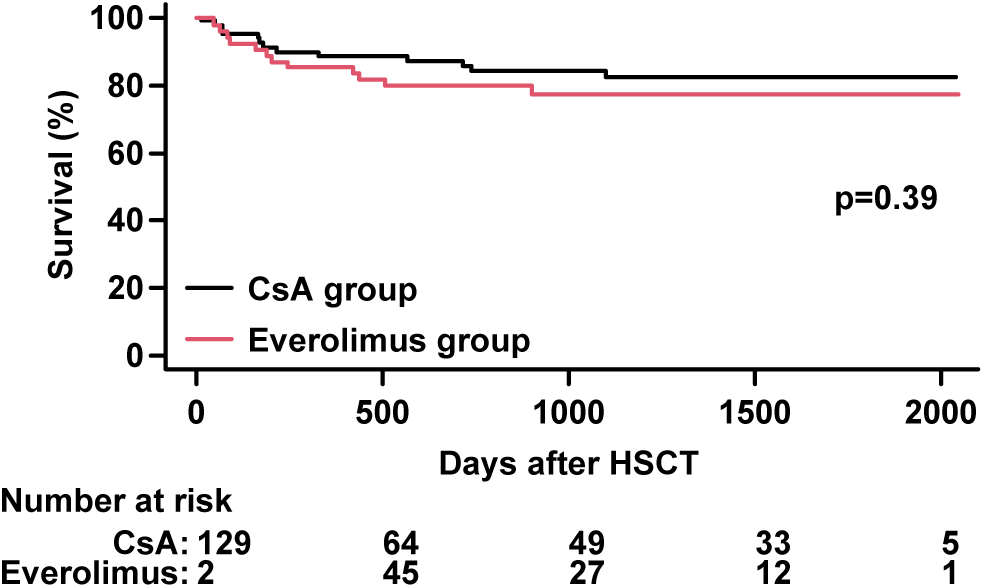
Overall survival (OS). The OS is shown by a Simon and Makuch plot to reflect the time dependent covariate of switch to everolimus. P-value is determined using the Mantel-Byar test.

**Figure 7.**
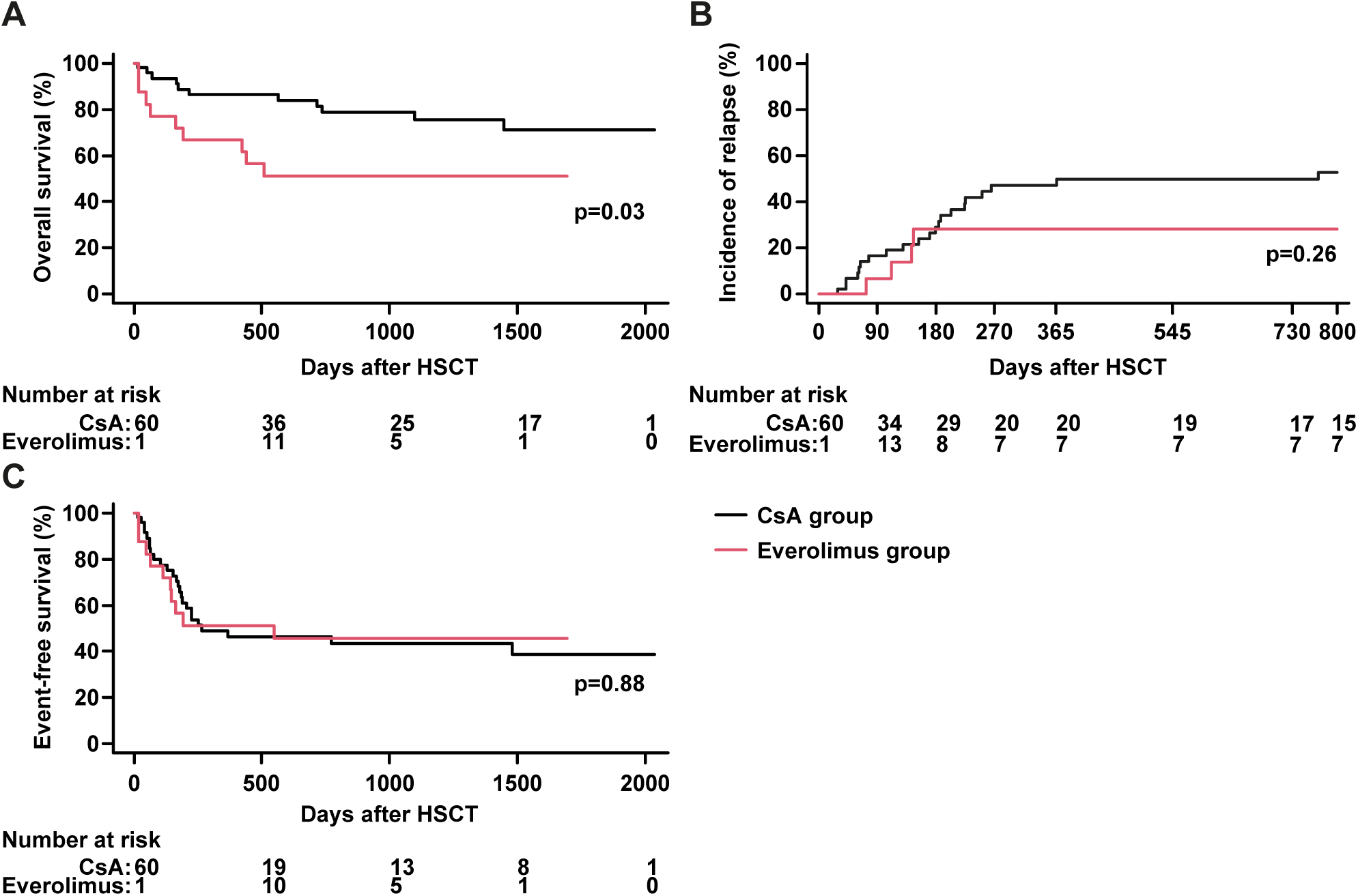
Overall survival (OS), incidence of relapse and event free survival (EFS) of patients with malignant underlying diseases. The patients of our cohort with malignant underlying diseases were selected and analyzed for their **(A)** OS, **(B)** incidence of relapse and **(C)** EFS with the time dependent covariate of switch to everolimus ± MMF. A and C show Simon and Makuch plots and B show a cumulative incidence using a competing events model. P-value is determined using the Mantel-Byar test.

## DISCUSSION

The aim of this retrospective analysis was to evaluate the feasibility of using everolimus in combination with or without MMF as GVHD prophylaxis when CNI had to be discontinued in a pediatric cohort of patients after their first allogeneic stem cell transplantation. The patients were treated at the Department of Pediatric Oncology and Hematology at the Charité – Universitätsmedizin Berlin between 2016 and 2019. The primary reasons for CNI discontinuation were nephrotoxicity and neurotoxicity. We show that, following the switch from ciclosporin A to everolimus ± MMF, retention parameters significantly decreased. The use of everolimus was not associated with a significantly higher overall incidence of acute or chronic GVHD. Despite AKI being considered a serious complication during post-transplant course, patients receiving everolimus exhibited a similar OS compared to the control cohort. Additionally, everolimus patients with a malignant underlying disease had a similar EFS compared to the control cohort. Subgroup analyses did not reveal any risk factors for the occurrence of GVHD or death. Here, we provide the first proof of concept, that everolimus in combination with MMF is a feasible immunosuppressive strategy in pediatric patients with AKI after allogeneic HSCT.

Our study significantly extends the existing data for the use of everolimus or mTOR inhibition in allogeneic HSCT. In adult transplantation, the addition of Sirolimus to standard CsA plus MTX for GVHD prophylaxis resulted in improved relapse-free and OS, with similar rates of advanced GVHD in a multicentre, randomised, phase 3 trial.^31^ Pidala *et al* demonstrated a lower incidence of acute and chronic GVHD in patients treated with sirolimus/tacrolimus than with MTX/tacrolimus in a randomized phase 2 trial.^32^ Everolimus was evaluated as GVHD prophylaxis in combination with tacrolimus in adults and appeared to be effective for the prevention of GVHD, however, the trial had to be terminated prematurely due to the development of sinusoidal obstruction syndrome in 25% of patients.^33^ In contrast to these results, we did not observe any life-threatening toxicity attributable to everolimus in our study. In three cases however, everolimus treatment had to be discontinued because of adverse events, such as stomatitis and interstitial pneumonitis. These adverse events were mild to moderate and in each case the ameliorated renal function allowed a re-conversion to CNI-treatment. A single-center phase I/II trial investigated the combination of everolimus and MMF as calcineurin inhibitor-free GVHD prophylaxis for 24 patients with hematologic malignancies and resulted in high rates of acute and chronic GHVD rates.^34^ The main difference to this study is that our patients converted from CNI-treatment to everolimus only after a median time of 22 days after HSCT. Only two patients received exclusively everolimus/MMF. The first (#53) suffered from severe aGVHD beginning on day 40 after HSCT and ultimately died due to severe cGVHD involving gut and lungs. The second patient (#15) is still alive three years after HSCT and shows no signs of cGVHD. Considering the literature and these cases, it appears that the very early stage after HSCT is critical for the development of acute and chronic GVHD. Therefore, it might be important to initiate CNI-based GVHD prophylaxis before safely incorporating everolimus/MMF. Supporting this hypothesis, other studies have identified the importance of sufficient CsA-levels during the first days after HSCT. Bianchi *et al* strongly recommend maintaining sufficient CsA levels during the initial 10 days after HSCT to reduce aGVHD.^35^ Additional studies demonstrate that a higher CsA starting dose of 5 mg/kg/day is independently associated with a lower risk for aGVHD^36^ or that lower CsA through levels lead to a higher incidence of grade II-IV aGVHD in the first four weeks after HSCT.^37^

Interestingly, we did not see a difference in OS between both patient groups, despite an increased mortality for pediatric patients with AKI being described in the literature.^7–10^ This observation might suggest that the conversion to everolimus/MMF could also have a beneficial effect on patient outcome, potentially counteracting the negative impact of AKI. One possible explanation might be a favorable effect on immune reconstitution. The role of everolimus and MMF in immune reconstitution, especially on CD4^+^ T cells, is not yet fully understood. Stable renal transplant recipients treated with an mTOR inhibitor, but not with CNI showed higher levels of circulating Tregs.^38^ In a murine GVHD model, treatment with rapamycin led to decreased activity of alloreactive conventional T cells, while regulatory T cells retained their immunosuppressive function, providing GVHD protection.^39^ In contrast to these results, Schaefer *et al* showed a protracted overall, regulatory and naïve CD4^+^ T cell reconstitution of adult patients that received everolimus/MMF after HSCT in a prospective single-center study compared with historical CNI-receiving controls.^34^ Furthermore, a beneficial effect could be explained by the direct anti-tumor efficacy of everolimus.^40,41^ The phosphatidylinositol-3-kinase (PI3K)/Akt and the mTOR signaling pathway, are hyperactivated in 50-80 % of AML patients^42^ and mTOR inhibition might be a possible therapeutic target. Everolimus monotherapy showed only limited therapeutic success.^43^ However, when combined with the hypomethylating agent azacitidine, everolimus led to promising results in OS and overall response rates in advanced AML.^44^ Also, for relapsed pediatric ALL, everolimus has been shown to be associated with favorable rates of complete remission and low end-reinduction MRD in a combination with a four-drug reinduction chemotherapy.^45^ In both combinations, everolimus seems to work as a sensitizer. However, whether this effect also contributes to the graft-versus-leukemia effect after HSCT remains to be shown.

With 131 patients, we were able to analyze a considerably large pediatric HSCT cohort. However, a larger group of patients would further increase the statistical interpretability. The underlying diseases and the used conditioning regimens varied substantially in our cohort, making it hard to identify specific subgroups that might benefit or be adversely affected by the conversion to everolimus/MMF. Along with the retrospective nature of the study, this diminishes the impact of our findings. In ongoing and future studies, it will be crucial to prospectively validate these results. Additionally, more detailed data on infectious complications and immune reconstitution should be collected.

In conclusion, our data represent the first clinical evidence that a conversion from CNI-based GVHD prophylaxis to everolimus/MMF during the clinical course after HSCT appears safe and feasible in pediatric patients across various underlying diseases. The earliest appropriate starting time of everolimus/MMF has yet to be defined but should probably not be during the first week post-HSCT. The duration of immunosuppressive treatment with everolimus/MMF strongly depends on the specific patient’s clinical course. Based on our data and in-house experience, we would generally recommend a duration of everolimus until day +180 after HSCT for benign underlying diseases and until day +80 for malignant underlying diseases in the absence of clinical evidence of acute or chronic GVHD.

## Data Availability

All data produced in the present study are available upon reasonable request to the authors.

## Acknowledgements

The authors thank Kathy Astrahantseff for editorial advice.

## Authorship contributions

FZ conceptualized the study, collected and analyzed data and prepared the manuscript. PL and JHS established the concept of CsA substitution by everolimus/MMF. ML, LO, JSK, AvS, SC, PH, AK, AE, & PL participated in the treatment of patients, in designing the study, discussing the data and reviewed the manuscript. BM and PG performed the cumulative incidence with competing events, gave input on statistical aspects and possible sources of bias and reviewed the manuscript. JHS co-conceptualized the study and reviewed the data, results and manuscript. All authors have read and agreed to the published version of the manuscript.

